# Internal Neurolysis for the Treatment of Trigeminal Neuralgia: Systematic Review

**DOI:** 10.1101/2021.05.25.21257791

**Authors:** Victor Sabourin, Pascal Lavergne, Jacob Mazza, Jeffrey Head, Fadi Al-Saiegh, Tony Stefanelli, Michael Karsy, James Evans

**Affiliations:** Department of Neurosurgery, Thomas Jefferson University Hospital, Philadelphia PA

**Keywords:** trigeminal neuralgia, internal neurolysis, facial pain

## Abstract

**Introduction:** Trigeminal neuralgia remains a challenging disease with significant debilitating symptoms and variable efficacy in terms of treatment options, namely microvascular decompression (MVD), stereotactic radiosurgery (SRS), and percutaneous rhizotomy. Internal neurolysis (IN) is an alternative treatment that may be provide patient benefit but has limited understanding. We performed a systematic review of IN treatment of trigeminal neuralgia.

**Methods:** Studies from 2000 to 2021 that assessed IN in trigeminal neuralgia were aggregated and independently reviewed. Weighted averages for demographics, outcomes and complications were generated.

**Results:** A total of 520 patients in 12 studies were identified with 384 who underwent IN (mean age 53.8 years, range 46-61.4 years). A mean follow-up time of 36.5 months (range 12-90 months) was seen. Preoperative symptoms were present for about 55.0 months before treatment and pain was predominantly in V2/3 (26.8%) followed by other distributions. An excellent to good outcome (Barrow Neurological Institute Pain Score [BNI-PS] I-III) was seen in 83.7% of patients (range 72-93.8%). Pain outcomes at 1 year were excellent in 58-78.4%, good or better in 77-93.75% and fair or better in 80-93.75% of patients. On average facial numbness following IN was seen in 96% of patients however at follow-up remained in only 1.75-10%. The vast majority of remaining numbness was not significantly distressing to patients. Subgroup comparisons of IN vs. recurrent MVD, IN vs. radiofrequency ablation, the impact of IN during the absence of vascular compression as well as IN with and without MVD were also evaluated.

**Conclusions:** IN represents a promising approach for surgical treatment of trigeminal neuralgia in the absence of vascular compression or in potential cases of recurrence. Complications were limited in general. Further study is required to evaluate the impact of IN via higher quality prospective studies.

## Introduction

Trigeminal neuralgia (TN) is a pain syndrome characterized by recurrent episodes of lancinating facial pain. The first line therapy for treatment is medical management, but many patients require surgery due to refractory symptoms or intolerable side effects from medication.^1,2^ The mainstay surgical treatment of TN is microvascular decompression (MVD) when neurovascular compression (NVC) is found.^2^

Although there is a strong association between NVC and TN, the pathophysiology is not completely understood.^1^ TN is known to occur and recur in the absence of NVC and many individuals with NVC do not manifest TN.^3–5^ One review by Lee *et al*. found that 4-89% of patients with TN do not have NVC, and in their own series found 28.8% of TN type I and 18.4% of TN type II patients had no NVC.^3^ Additionally, for MVDs there is a difference in outcome based on severity of NVC, and whether or not the compression is arterial or venous.^2,5–11^ Furthermore, although high resolution MRI/MRA are reliable tests in verifying NVC with a sensitivity of 96% for TN type I + II and a specificity of 90% for TN I and 66% for TN II, there are still false positives with no NVC seen at time of surgery.^3^

The subgroup of patients presenting with TN in the absence of NVC or with low grade arterial compression or venous compression are harder to manage. In these cases, MVD is either not possible or associated with a higher rate of treatment-failure, which prompts the need for other treatment options. Percutaneous radiofrequency rhizotomy (RF), glycerol rhizotomy, balloon compression, stereotactic radiosurgery (SRS), and partial sensory rhizotomy (PSR) have been the traditional second line surgical therapies for this patient population).^1,12–22^

However, recently another surgical option, Internal Neurolysis (IN), has emerged as an attempt to provide long-term pain relief to those refractory patients. Internal Neurolysis, also known as “nerve-combing”, is the process of microsurgical parallel dissection of the cisternal portion of the trigeminal nerve into multiple nerve fasicles.^1^ Although the first reports seemed promising, the efficacy, durability, and complication pattern remain to be fully defined.^23^ This manuscript provides a systematic review of available literature on the efficacy of IN for treating patients with TN.

## Methods

The Preferred Reporting Items for Systematic Reviews and Meta-Analyses (PRISMA) guidelines were followed for reporting our systematic review.^24^

### Eligibility Criteria

We included studies reporting internal neurolysis of the trigeminal nerve as a surgical treatment for trigeminal neuralgia (Figure 1). Inclusion criteria were as follows: 1) Articles which analyzed outcomes of IN with or without a comparative group, 2) Articles had to report pain control outcome with at least one year follow-up, 3) Study designs could include randomized controlled trials, prospective, or retrospective cohort study, 4) Study in English language, and 5) Study published between 2000-2021. Series that did not differentiate the results of IN from other treatments were excluded. Studies were required to have at least 1 year of follow-up to assess the durability of IN. The search was constrained to the years 2000-2021 to ensure that collected data analyzed would be recent and to also help prevent potentially outlier data related to evolving surgical technique.

**Figure 1:**
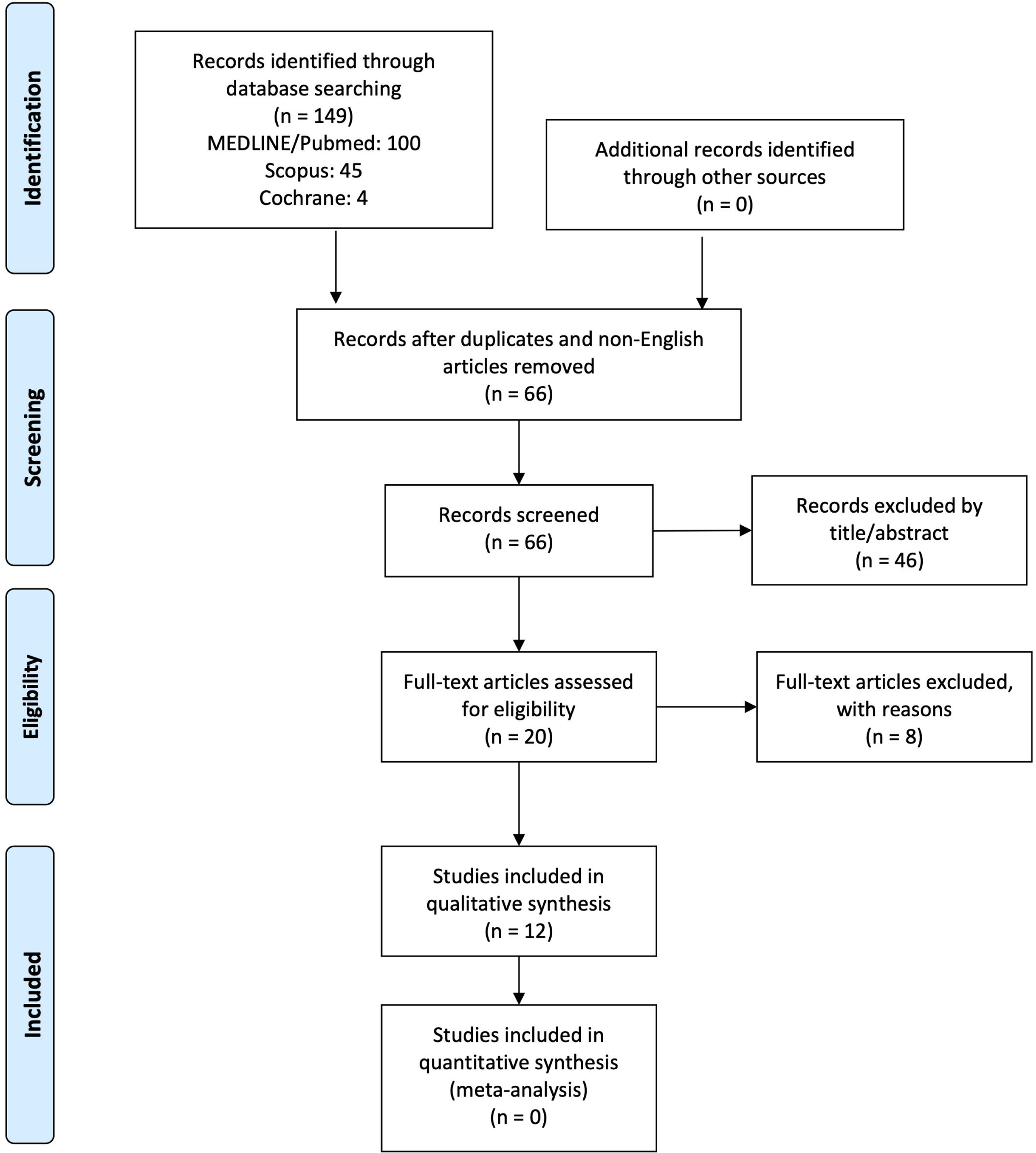
PRISMA 2009 Flow Diagram.

### Information Sources and Search Strategy

MEDLINE (PubMed), Scopus, and Cochrane databases were queried from 01/01/2000 to 4/8/2021. Ongoing studies were searched in the ClinicalTrials.gov registry, the controlled-trials.com registry, and the Trials Central databases. References from the included studies were screened for additional references. The search strategy included the following terms as well as appropriate indexation term: “Trigeminal Neuralgia, Neurolysis”, “Trigeminal Neuralgia, Internal Neurolysis”, and “Trigeminal Neuralgia, Microvascular Decompression, Neurolysis”. There was no restriction on study design or outcomes in the search strategy.

### Study Selection and data extraction

Three independent reviewers screened titles and abstract after accounting for duplicates and eliminating non-English articles. Full text was retrieved for included articles after primary screening. These articles as well as their citations were fully reviewed for eligibility. If there were any discrepancy between any of the reviewers, articles were included by consensus or with the help of a third reviewer.

The following data was searched for and extracted from the included articles: Patient number, sex, TN type, treatment type, age, length of follow-up, pre-operative symptom duration, TN facial distribution, previous surgical history, pain outcomes, hypesthesia outcomes, rates of recurrence, complications, Barrow Neurological Institute Pain Scale (BNI-PS) – a pain scoring system, Barrow Neurological Institute Hypesthesia Scale (BNI-HS) – a hypesthesia scoring system, University of San Francisco Pain Score (UCSF Pain Scale) – a pain scoring system, Numerical Pain Rating Scale – a basic scoring system which can be used to assess different factors such as pain and quality of life.

### Outcomes measures

Primary outcomes were post-operative pain score as defined by the BNI-PS or UCSF pain score. Outcome were stratified as either excellent (BNI-PS 1 or UCSF excellent), good (BNI-PS I/II or UCSF excellent) or fair (BNI-PS: I-III and UCSF pain score: excellent/good). Secondary outcomes included recurrence rate and complication.

### Risk of Bias in Individual Studies

Specific analysis of bias risk was not performed as these studies were all retrospective, non-randomized trials without blinded assessment of outcomes. Where relevant, missing data are reported in the summary tables. No specific method was used to assess risk of bias in individual studies.

### Summary Measures

Weighted averages for patients who underwent IN were generated for continuous variables, including patient demographics, outcomes, and complications, when available. Averages for outcomes used the study definitions of a good outcome or otherwise considered BNI-PS I-III (Excellent to Good) as a good outcome. The average rates of complications, such as the number of patients reporting facial numbness, at the last known follow-up was noted. Subgroups of patients with only IN were also analyzed for outcomes and complications. For studies that did not report mean ages, the median age was used. Variable ranges are reported when available.

## Results

### Patient Demographics

A total of 520 patients were included in the 12 studies,^1,23,25–34^ 384 (73.8%) of whom underwent IN (Table 1). Mean/median age for all studies was 53.8 years old (range 46-61.4 years) and mean/median follow-up time was 36.5 months (range 12-90 months). The pre-operative symptom duration was included in 8 studies and averaged 55.0 months (range 40.4 – 70.8 months). When given, the TN type as well as facial distributions were entered. The average TN distribution was most common for V2/3 (26.8%) followed by V1-2 (16.1%), V1-3 (16.1%), and V3 (15.6%). A total of 5 studies reported previously attempted surgical treatments in their patient populations.

**Table 1:**
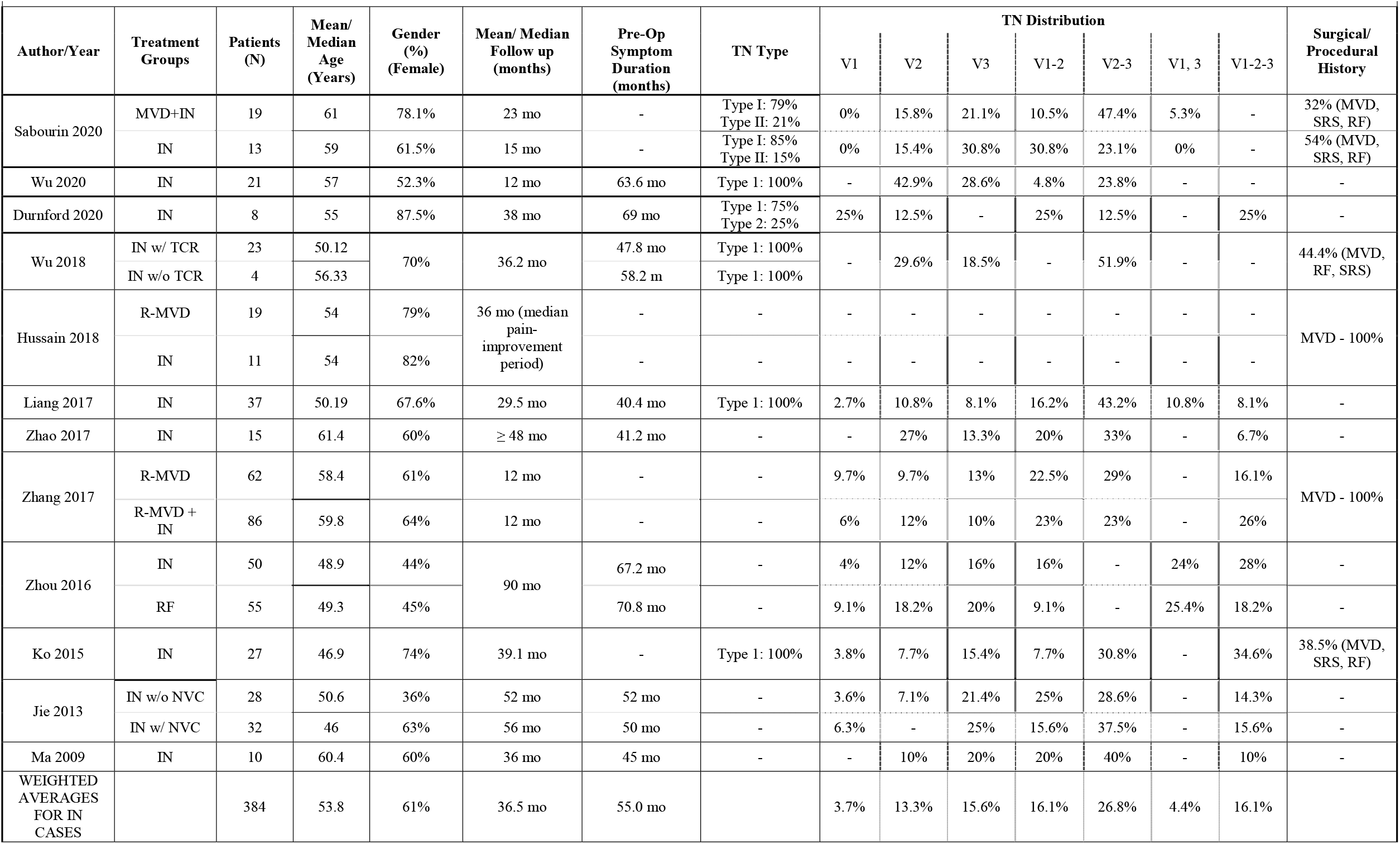
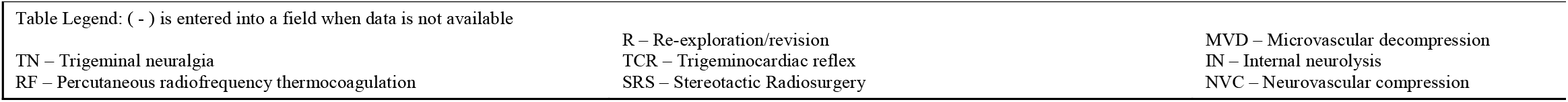
Study Demographics and Patient Characteristics.

| Author/Year | Treatment Groups | Patients (N) | Mean/ Median Age (Years) | Gender (%) (Female) | Mean/ Median Follow up (months) | Pre-Op Symptom Duration (months) | TN Type | TN Distribution |  |  |  |  |  |  | Surgical/ Procedural History |
| --- | --- | --- | --- | --- | --- | --- | --- | --- | --- | --- | --- | --- | --- | --- | --- |
|  |  |  |  |  |  |  |  | V1 | V2 | V3 | V1-2 | V2-3 | V1, 3 | V1-2-3 |  |
| Sabourin 2020 | MVD+IN | 19 | 61 | 78.1% | 23 mo | - | Type I: 79%<br>Type II: 21% | 0% | 15.8% | 21.1% | 10.5% | 47.4% | 5.3% | - | 32% (MVD, SRS, RF) |
|  | IN | 13 | 59 | 61.5% | 15 mo | - | Type I: 85%<br>Type II: 15% | 0% | 15.4% | 30.8% | 30.8% | 23.1% | 0% | - | 54% (MVD, SRS, RF) |
| Wu 2020 | IN | 21 | 57 | 52.3% | 12 mo | 63.6 mo | Type 1: 100% | - | 42.9% | 28.6% | 4.8% | 23.8% | - | - | - |
| Durnford 2020 | IN | 8 | 55 | 87.5% | 38 mo | 69 mo | Type 1: 75%<br>Type 2: 25% | 25% | 12.5% | - | 25% | 12.5% | - | 25% | - |
| Wu 2018 | IN w/ TCR | 23 | 50.12 | 70% | 36.2 mo | 47.8 mo | Type 1: 100% | - | 29.6% | 18.5% | - | 51.9% | - | - | 44.4% (MVD, RF, SRS) |
|  | IN w/o TCR | 4 | 56.33 |  |  | 58.2 m | Type 1: 100% |  |  |  |  |  |  |  |  |
| Hussain 2018 | R-MVD | 19 | 54 | 79% | 36 mo (median pain-improvement period) | - | - | - | - | - | - | - | - | - | MVD - 100% |
|  | IN | 11 | 54 | 82% |  | - | - | - | - | - | - | - | - | - |  |
| Liang 2017 | IN | 37 | 50.19 | 67.6% | 29.5 mo | 40.4 mo | Type 1: 100% | 2.7% | 10.8% | 8.1% | 16.2% | 43.2% | 10.8% | 8.1% | - |
| Zhao 2017 | IN | 15 | 61.4 | 60% | ≥ 48 mo | 41.2 mo | - | - | 27% | 13.3% | 20% | 33% | - | 6.7% | - |
| Zhang 2017 | R-MVD | 62 | 58.4 | 61% | 12 mo | - | - | 9.7% | 9.7% | 13% | 22.5% | 29% | - | 16.1% | MVD - 100% |
|  | R-MVD + IN | 86 | 59.8 | 64% | 12 mo | - | - | 6% | 12% | 10% | 23% | 23% | - | 26% |  |
| Zhou 2016 | IN | 50 | 48.9 | 44% | 90 mo | 67.2 mo | - | 4% | 12% | 16% | 16% | - | 24% | 28% | - |
|  | RF | 55 | 49.3 | 45% |  | 70.8 mo | - | 9.1% | 18.2% | 20% | 9.1% | - | 25.4% | 18.2% | - |
| Ko 2015 | IN | 27 | 46.9 | 74% | 39.1 mo | - | Type 1: 100% | 3.8% | 7.7% | 15.4% | 7.7% | 30.8% | - | 34.6% | 38.5% (MVD, SRS, RF) |
| Jie 2013 | IN w/o NVC | 28 | 50.6 | 36% | 52 mo | 52 mo | - | 3.6% | 7.1% | 21.4% | 25% | 28.6% | - | 14.3% | - |
|  | IN w/ NVC | 32 | 46 | 63% | 56 mo | 50 mo | - | 6.3% | - | 25% | 15.6% | 37.5% | - | 15.6% | - |
| Ma 2009 | IN | 10 | 60.4 | 60% | 36 mo | 45 mo | - | - | 10% | 20% | 20% | 40% | - | 10% | - |
| WEIGHTED AVERAGES FOR IN CASES |  | 384 | 53.8 | 61% | 36.5 mo | 55.0 mo |  | 3.7% | 13.3% | 15.6% | 16.1% | 26.8% | 4.4% | 16.1% |  |

|  |  |  |
| --- | --- | --- |
| Table Legend: ( - ) is entered into a field when data is not available |  |  |
| TN – Trigeminal neuralgia | R – Re-exploration/revision | MVD – Microvascular decompression |
| RF – Percutaneous radiofrequency thermocoagulation | TCR – Trigemino-cardiac reflex | IN – Internal neurolysis |
|  | SRS – Stereotactic Radiosurgery | NVC – Neurovascular compression |

### Overall Pain Outcomes

Overall excellent to good outcomes (BNI-PS I-III) were seen in 83.7% of patients (range 72-93.8%) (Table 2). Good outcomes did show a slight decrease over follow-up time. The immediate post-operative results were excellent in 85-94.6%, good or better in 96-100% and fair or better in 96-100% of patients. When looking the 1-year post-op pain outcomes, the rates ranged as follows: excellent in 58-78.4%, good or better in 77-93.75% and fair or better in 80-93.75% of patients. The primary outcome for all studies irrespective of follow-up time was excellent in 47-82.1%, good or better in 62.5-87.1% and fair or better in 80-100% of patients. Several studies were likely underestimated outcomes since only a BNI PS I-II could be obtained.

**Table 2:**
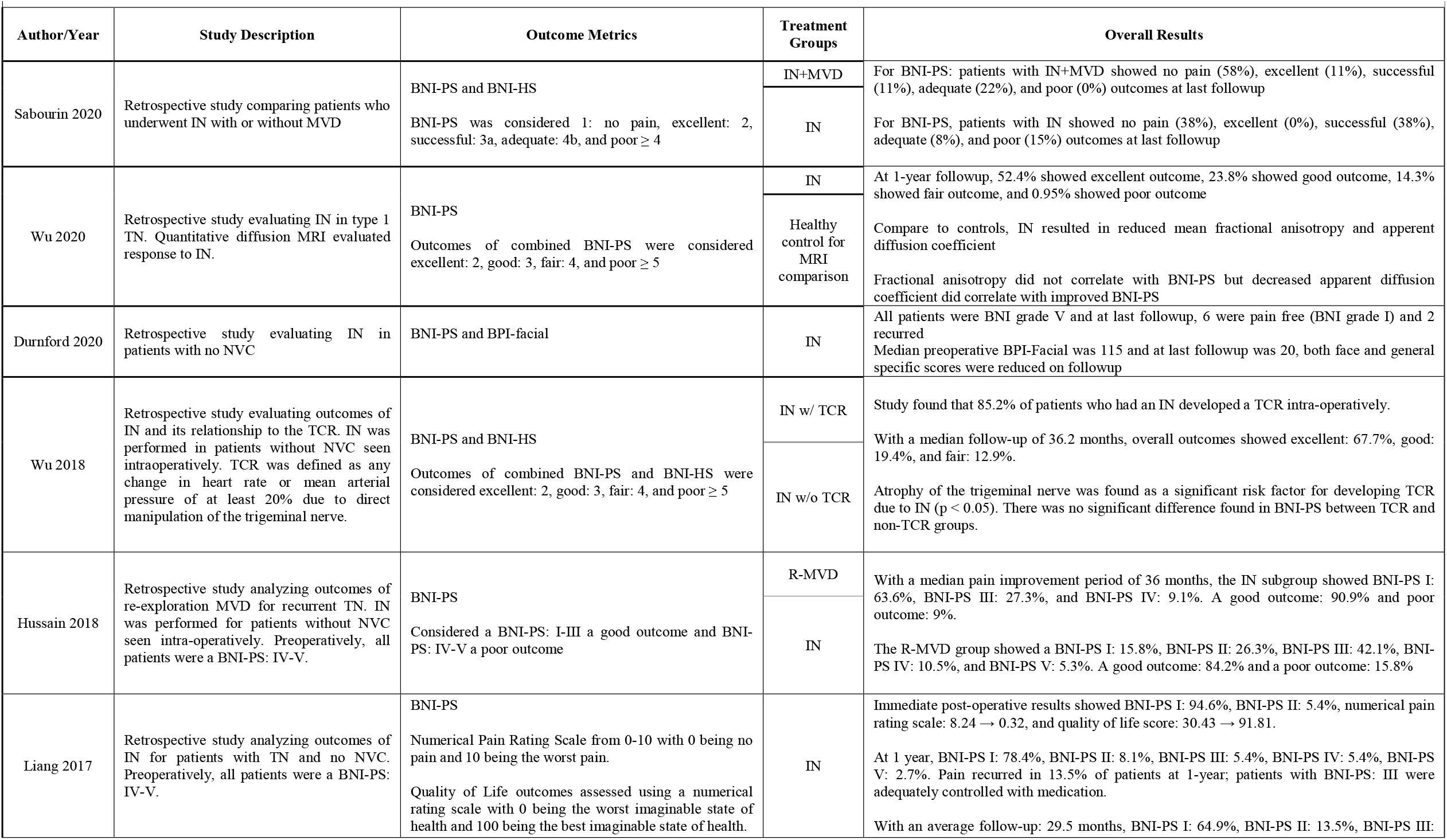
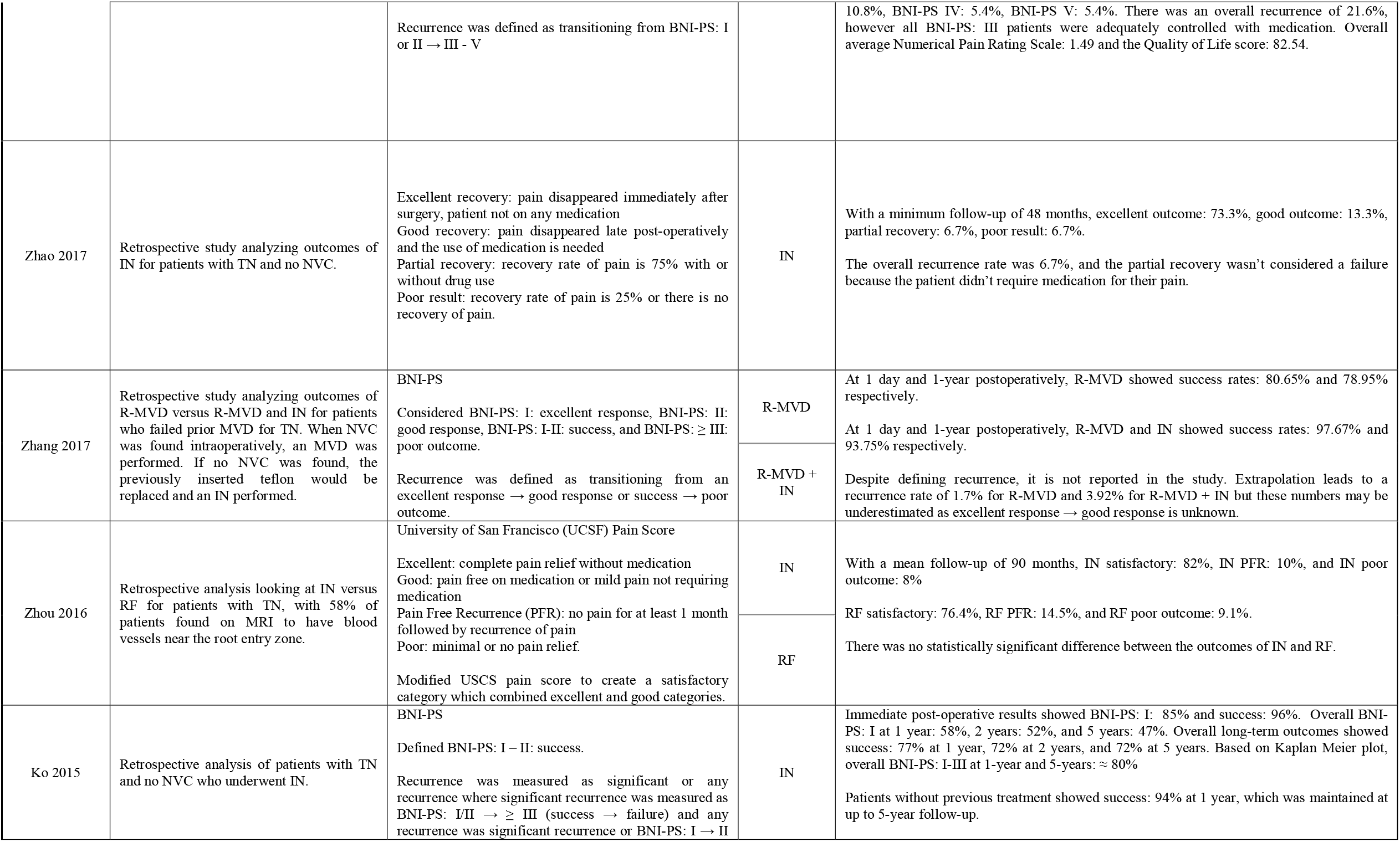
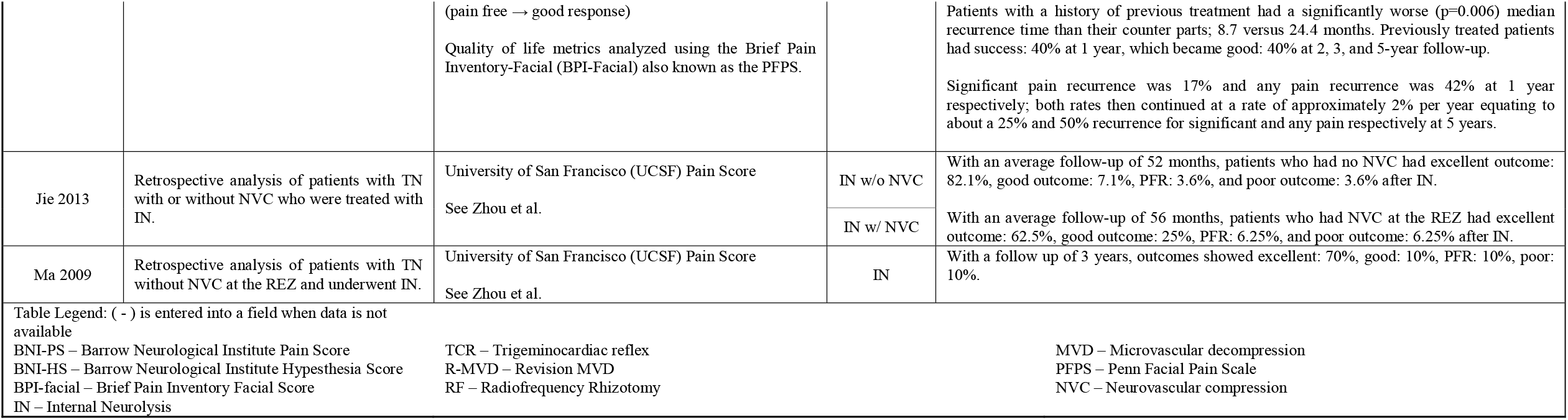
Study Description with Outcome Metrics and Treatment Outcomes.

### Recurrence

The 1-year recurrence rates of any pain including a transition of BNI-PS from I to II ranged from 3.92-42% and the overall rate was 3.6-50%. However, when considering only significant recurrence of pain, defined as BNI-PS I/II to III-V, the 1-year recurrence rates ranged from: 3.92-17%, and the overall recurrence rates ranged from: 3.6-25%.

### IN Outcomes with NVC present

Three studies included data on IN for patients with NVC. In Zhou et al.’s patient population, 58% of the patients were found on MRI to have a blood vessel near the REZ and showed an overall satisfactory result of 82% after IN as defined by the study. Additionally, Jie et. al compared the results of IN in patients with and without NVC and found that with over 4 years of follow-up, patients without NVC had a 19.6% higher rate of excellent outcome as well as overall lower rates of recurrence and poor outcomes on the UCSF Pain Score.^27^ Sabourin et al. showed good outcomes were achieved in 80% of patients with IN and NVC compared with 76% in patients with IN alone, which suggests a very limited difference between these two groups of patients.^29^

### Re-Exploration MVD with and without IN

Two studies specifically evaluated re-exploration MVD with and without IN for recurrent TN. Hussain et al. found that patients who underwent an IN (84.2%) had similar overall rates of a good outcome compared with re-exploration MVD (90.9%), but patients who underwent an IN had a 47.8% higher rate of BNI-PS I outcomes. Zhang et al. found that at 1 year, patients who had undergone a revision MVD and IN had a 14.8% higher rate of success as defined by the study compared to patients who underwent revision MVD alone.^26,32^

### IN Outcomes in Relation to Previous Treatment(s) for TN

There were 5 studies which reported a patient population with previous treatment including MVD, stereotactic radiosurgery (SRS) and radiofrequency (RF) ablation (Table 1). A range of 32-100% of patients underwent previous treatments. Ko et al. stratified patients with prior MVD and found that patients with a history of previous treatment did significantly worse than their counter parts.^1^ Patients with a history of previous treatment showed 40% success (BNI-PS I/II) at 1-year which became 40% good outcomes (BNI ≥III) at 2, 3, and 5 year follow-up. Patients without a history of previous treatment showed 94% success at 1-year, which was maintained at 2, 3, and 5 years of follow-up. The median time to recurrence for patients with a history of previous procedure was 8.7 months while for those without was 24.4 months.

### IN compared to RF

One study compared IN with RF and found a trend toward IN having higher satisfactory rates, lower recurrence rates, and lower poor outcomes when compared to RF. However, results were not statistically significant.^34^

### Complications

The primary complication of IN is facial numbness (e.g., hypesthesia, hypoesthesia) (Table 3). Rates of facial numbness following IN were as high as 96% in the immediate post-operative period, but at long-term follow-up, numbness decreased to about 38.8% on average. However, studies varied significantly on follow-up length and when complications occurred. In addition, long-term numbness was often mild and non-distressing in the majority of studies with only 1.75-10% seen in selected studies that fully reported complications at last follow-up. As IN is a technique involving direct manipulation of the trigeminal nerve, other important complications to consider are corneal hypesthesia, corneal ulcer, loss of corneal reflex, and anesthesia dolorosa. The overall rate of corneal hypesthesia and ulcer within the review was 1.2%, and 1 case of anesthesia dolorosa was reported (0.31%). There were several other surgical complications mentioned in the articles reviewed including, facial nerve dysfunction, CSF leak, and meningitis, however these complications were related more to the surgical approach.

**Table 3:**
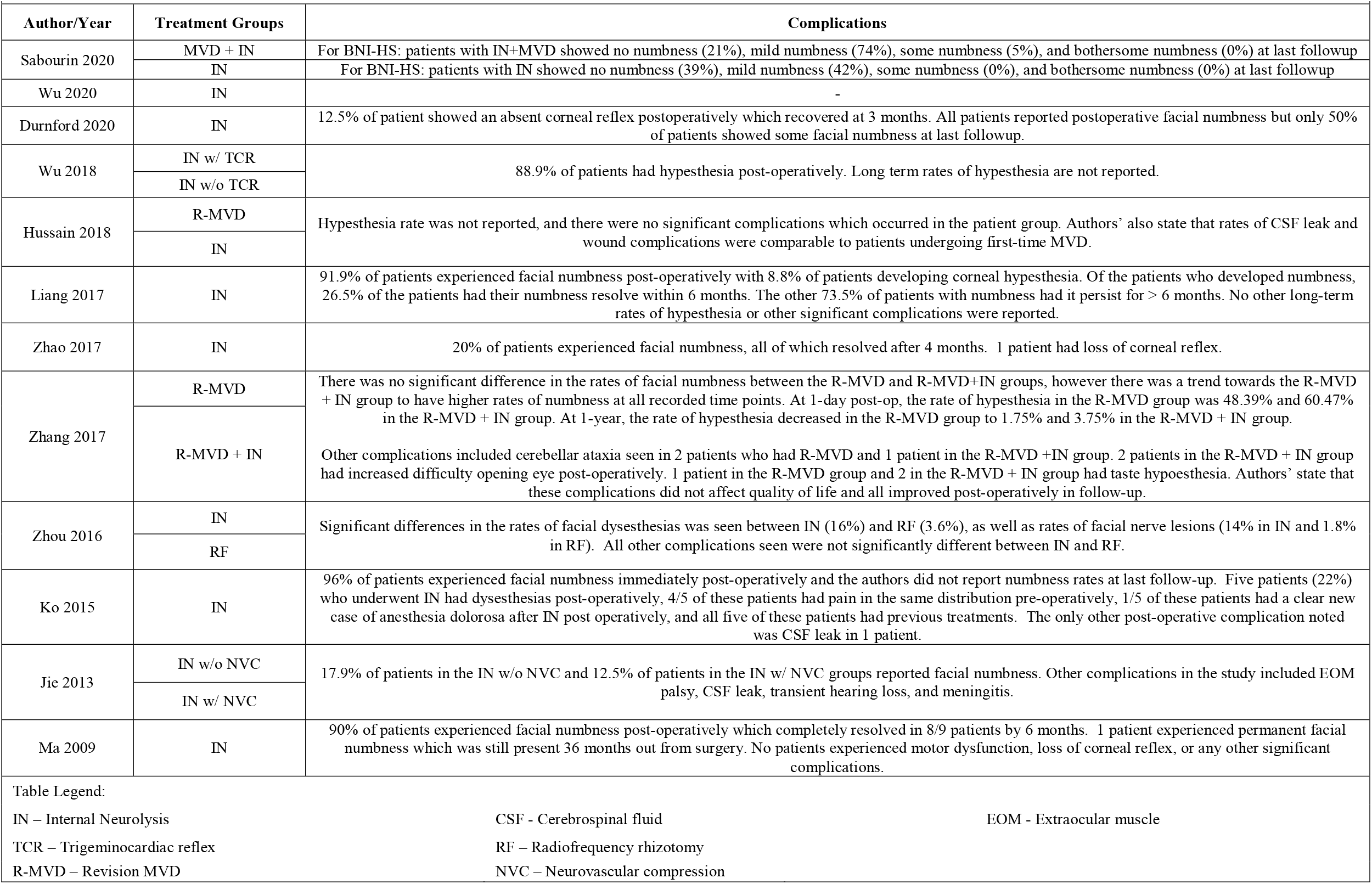
Complications.

## Discussion

Our systematic review is the first one to address the surgical result of posterior fossa exploration with IN of the trigeminal nerve for trigeminal neuralgia. Our results shows that the overall excellent to good outcomes (BNI-PS I-III) were seen on average in 83.7% of patients who underwent IN. Recurrence rates for clinically significant changes in pain (BNI-PS I/II to III-V) ranged from 3.6-25%. Several studies showed improved BNI-PS I outcomes after IN compared with re-exploration MVD alone and patients without any history of prior TN surgical treatment seemed to fare better. Some, but not all studies, showed that patients with re-exploration MVD had better results if an IN was performed. However, heterogeneity between studies did not allow for clear answers regarding the concomitant role of MVD and IN, as well as role of IN alone in recurrent disease. The consequences of trigeminal nerve manipulation was high rates of post-operative facial hypesthesia seen in up to 96% of patients in some studies. Numbness was seen on average in 38.8% of patients after varying lengths of follow-up. Only 1.75-10% of patients still had noticeable facial numbness at last follow-up. The major complication of corneal ulceration or anesthesia dolorosa were rare with each seen in single cases.

### IN versus MVD outcomes

MVD for type I TN remains a durable surgical treatment option and comparison of IN outcomes are limited. IN is considered in cases without NVC on imaging or direct surgical observation. Excellent results (pain free) with initial MVD alone can be seen in 82% of patients immediately postoperatively, 75% at 1 year, and 64% at 10 years.^2^ The pain control rates of IN, although good, are not as robust as those seen with MVD. Overall good outcomes were seen in 83.7% of cases with a mean follow-up of 36.5 months. The results of this study suggest varying follow-up length of available studies confounds the definitive comparison of outcomes with MVD vs. MVD with IN. Sindou et al. showed that at 1 and 15 years respectively, BNI-PS I cure rates considering NVC grade were 96.6% and 88.1% (Grade III: adhesion), 90.2% and 78.3% (Grade II: Touching & Indentation), and 83.3% and 58.3% (Grade I: Touching), respectively.^6,10^ Venous compression also appears to have decreased pain control rates, showing decreased BNI-PS I rates of 8.1% and 14.3% at 1 and 15 years compared to arterial compression, respectively.^6^ These longer follow-up lengths are simply not available for IN and especially for first-time IN treatments.

Results after MVD for Grade III compression appear superior to IN. But MVD in lower grade NVC or venous compression appear comparable to results seen for IN. Two studies reported IN on patients with NVC, however only one stratified the results and the severity of the NVC was not specified. Despite the fact that most patients undergoing an IN have no NVC, the potential for adding IN in milder NVC cases with recurrent pain may be a promising option for patients.

Another indication of IN is for recurrent pain after MVD. Zhang et al. reported 1-year success rates of 93.75% after revision MVD with IN.^32^ Hussain et al. suggested similar good outcomes between revision MVD and IN at last follow-up (90.9% vs. 84.2%).^26^ Rates of excellent results after revision MVD without IN appear to be 50-60%, 40-50% and around 42%, at 1-, 5-, and 10-years, respectively.^2^ This shows that re-exploration with IN should be considered for pain recurrence after MVD. Whether this strategy is better that radiosurgery or percutaneous rhizotomy remained to be determined.

### IN vs radiosurgery

A systematic review of SRS for TN showed that average rates of initial pain freedom with or without medications after a latency period were: GKS: 84.8%, LINAC: 87.3%, and Cyberknife: 79% without any significant difference between the radiation modalities.^15^ Average rates of pain freedom after SRS without medication use were: GKS: 53.1%, LINAC: 49.3%, and Cyberknife: 56.3%, again without any significant difference between treatment modalities.^15^ The review also found 2 studies reporting rates of pain freedom without medication at 10 years were 30% and 45.3% and that previous surgery was a negative predictor of pain relief after SRS.^15,19,35^ Within the limits of the data for IN which exists, IN appears to have a higher success rate when compared to SRS in short-term follow-up and remains to be seen for long-term follow-up.

### IN versus Percutaneous Procedures

Many retrospective cohort studies have described the success rates of the different percutaneous procedures. Success usually ranges between 90-97% pain relief in the immediate post-operative period, but with recurrence rate as high as 75% at long-term follow-up^20,26,28,29^. When comparing the different techniques, RF appears to provide the best pain relief among the percutaneous procedures, but also showed a potentially higher complication profile.^21^ One review found a trend of RF showing a higher rates of anesthesia dolorosa as a complication compared to glycerol rhizotomy or balloon compression.^21^

One study in our systematic review compared the results of RF versus IN. Zhou et al. found that with an average follow-up of 90 months, there was a trend towards IN producing higher rates of satisfactory results as well as lower recurrence rates and lower poor outcomes as defined by the study.^34^ However, because of a significantly higher complication rate seen with IN compared to RF and a non-significant difference in treatment outcomes, the authors concluded that RF was the preferred procedure. The higher complications seen in the IN group were facial dysesthesia in 16% of patients and facial nerve injury in 14% of patients.^34^ This rate of facial nerve injury with posterior fossa exploration is higher than most published studies that usually range between 0.5-3%^2^. Furthermore, in our systematic review, the rate of facial dysesthesia was around 4%. When considering the results of these different studies, IN appears to be as effective and potentially more effective than percutaneous procedures. It seems particularly indicated in patients without NVC and any prior TN treatment.

### IN versus PSR

PSR has been used as a surgical option to treat TN prior to MVD.^36^ It is an alternative to MVD with absent intraoperative NVC, cases with venous compression, and during revision surgery for failed prior MVD.^17,37–39^ One series^17^ that used these criteria for PSR found excellent, good, and poor results in 48%, 22%, and 30% of patients, respectively. In their series, 42% of patients had a prior treatment for TN, 76% had no NVC intraoperatively and 23% were found to have NVC and underwent either isolated PSR or PSR with MVD. Results for patients with no prior posterior fossa surgery were excellent in 64% at 1 year and excellent in 55-60% at 5 years. On the other hand, patients with prior posterior fossa surgery showed worsened results – excellent in 38% at 1 month and 1 year, as well as excellent in 10-15% at 5 years.^17^

As for IN, patients with previous surgery fared worse with PSR. However, the overall excellent result of 48% for PSR at 5 years fell within the lower range seen for IN and the outcome of 55-60% excellent results after PSR in patients without previous surgery is much lower than the 94% excellent results seen with IN in patients without previous treatment.^1^

### Complications

The complications found in this systematic review were comparable to those found in other treatments for TN except for facial numbness. It is an expected complication of IN that increased manipulation of the trigeminal nerve results in increased numbness. The results here suggested significant recovery of numbness over time in the IN patient population, from 96% of patients at post-op to 1-10% at last follow-up. Furthermore, these studies suggested that painful numbness was fairly low. Further studies are warranted to better define the post-op incidence, the distribution, and the recovery of the facial hypesthesia. The relation between post-operative facial numbness and long-term pain relief needs to be further established. Major complications related to the trigeminal nerve dissection, including corneal ulceration and anesthesia dolorosa, have only been rarely report in the included studies. The rest of the complication profile appears similar to MVD.

### Limitations

Although it is the first systematic review on the use of trigeminal nerve IN, there are several limitations. All studies included in the review have a retrospective design with unblinded assessment of outcomes and were thus subject to a high risk of bias. There was also a high degree of heterogeneity in the patient populations within the different studies, which may create variability in the pain relief outcomes. Additionally, the follow-up times for studies in this review remain relatively short in comparison to published studies on other treatment modalities for TN, which also makes it difficult to compare results.

## Conclusions

IN for TN may be effective in providing pain relief for patient. The results of our review suggest that IN might be as effective as MVD in patient with low grade NVC. Furthermore, in patient with no NVC or for recurrence after MVD, IN seems to provide at least similar if not better short-term outcomes than the other surgical options. These patient population are notably harder to treat with higher TN recurrence rate. IN is another treatment option that might allow better long-term pain relief than SRS or percutaneous treatment options. The tradeoff appears to be an increased in the immediate facial hypoesthesia which seems to improve in the majority of patients. Future studies, ideally with prospective designs, randomized trials and registries are required to better define the long-term pain relief, ideal patient population, and complication profile of IN.

## Data Availability

The data are available in the primary papers from where this review was acquired.

## Notes

### Competing Interest Statement

The authors have declared no competing interest.

### Funding Statement

No external funding was received for this study

### Author Declarations

IRB excemption, review paper

## References

1. Ko AL, Ozpinar A, Lee A, Raslan AM, McCartney S, Burchiel KJ. Long-term efficacy and safety of internal neurolysis for trigeminal neuralgia without neurovascular compression. Journal of neurosurgery. 2015;122(5):1048–1057.

2. Barker FG, 2nd, Jannetta PJ, Bissonette DJ, Larkins MV, Jho HD. The long-term outcome of microvascular decompression for trigeminal neuralgia. The New England journal of medicine. 1996;334(17):1077–1083.

3. Lee A, McCartney S, Burbidge C, Raslan AM, Burchiel KJ. Trigeminal neuralgia occurs and recurs in the absence of neurovascular compression. J Neurosurg. 2014;120(5):1048–1054.

4. Miller JP, Acar F, Hamilton BE, Burchiel KJ. Radiographic evaluation of trigeminal neurovascular compression in patients with and without trigeminal neuralgia. Journal of neurosurgery. 2009;110(4):627–632.

5. Leidinger A, Munoz-Hernandez F, Molet-Teixido J. Absence of neurovascular conflict during microvascular decompression while treating essential trigeminal neuralgia. How to proceed? Systematic review of literature. Neurocirugia. 2018;29(3):131–137.

6. Sindou M, Leston J, Decullier E, Chapuis F. Microvascular decompression for primary trigeminal neuralgia: long-term effectiveness and prognostic factors in a series of 362 consecutive patients with clear-cut neurovascular conflicts who underwent pure decompression. Journal of neurosurgery. 2007;107(6):1144–1153.

7. Burchiel KJ, Clarke H, Haglund M, Loeser JD. Long-term efficacy of microvascular decompression in trigeminal neuralgia. Journal of neurosurgery. 1988;69(1):35–38.

8. Kolluri S, Heros RC. Microvascular decompression for trigeminal neuralgia. A five-year follow-up study. Surgical neurology. 1984;22(3):235–240.

9. Toda H, Iwasaki K, Yoshimoto N, et al. Bridging veins and veins of the brainstem in microvascular decompression surgery for trigeminal neuralgia and hemifacial spasm. Neurosurgical focus. 2018;45(1):E2.

10. Leal PR, Hermier M, Froment JC, Souza MA, Cristino-Filho G, Sindou M. Preoperative demonstration of the neurovascular compression characteristics with special emphasis on the degree of compression, using high-resolution magnetic resonance imaging: a prospective study, with comparison to surgical findings, in 100 consecutive patients who underwent microvascular decompression for trigeminal neuralgia. Acta neurochirurgica. 2010;152(5):817–825.

11. Hardaway FA, Gustafsson HC, Holste K, Burchiel KJ, Raslan AM. A novel scoring system as a preoperative predictor for pain-free survival after microsurgery for trigeminal neuralgia. Journal of neurosurgery. 2019:1–8.

12. Wu CY, Meng FG, Xu SJ, Liu YG, Wang HW. Selective percutaneous radiofrequency thermocoagulation in the treatment of trigeminal neuralgia: report on 1860 cases. Chinese medical journal. 2004;117(3):467–470.

13. Kondziolka D, Perez B, Flickinger JC, Habeck M, Lunsford LD. Gamma knife radiosurgery for trigeminal neuralgia: results and expectations. Archives of neurology. 1998;55(12):1524–1529.

14. Lopez BC, Hamlyn PJ, Zakrzewska JM. Stereotactic radiosurgery for primary trigeminal neuralgia: state of the evidence and recommendations for future reports. Journal of neurology, neurosurgery, and psychiatry. 2004;75(7):1019–1024.

15. Tuleasca C, Regis J, Sahgal A, et al. Stereotactic radiosurgery for trigeminal neuralgia: a systematic review. Journal of neurosurgery. 2018;130(3):733–757.

16. Kondziolka D, Lunsford LD. Percutaneous retrogasserian glycerol rhizotomy for trigeminal neuralgia: technique and expectations. Neurosurgical focus. 2005;18(5):E7.

17. Young JN, Wilkins RH. Partial sensory trigeminal rhizotomy at the pons for trigeminal neuralgia. Journal of neurosurgery. 1993;79(5):680–687.

18. Wu H, Zhou J, Chen J, Gu Y, Shi L, Ni H. Therapeutic efficacy and safety of radiofrequency ablation for the treatment of trigeminal neuralgia: a systematic review and meta-analysis. Journal of pain research. 2019;12:423–441.

19. Kondziolka D, Zorro O, Lobato-Polo J, et al. Gamma Knife stereotactic radiosurgery for idiopathic trigeminal neuralgia. Journal of neurosurgery. 2010;112(4):758–765.

20. Cheng JS, Lim DA, Chang EF, Barbaro NM. A review of percutaneous treatments for trigeminal neuralgia. Neurosurgery. 2014;10 Suppl 1:25-33; discussion 33.

21. Texakalidis P, Xenos D, Tora MS, Wetzel JS, Boulis NM. Comparative safety and efficacy of percutaneous approaches for the treatment of trigeminal neuralgia: A systematic review and meta-analysis. Clin Neurol Neurosurg. 2019;182:112–122.

22. Xu Z, Schlesinger D, Moldovan K, et al. Impact of target location on the response of trigeminal neuralgia to stereotactic radiosurgery. Journal of neurosurgery. 2014;120(3):716–724.

23. Ma Z, Li M. “Nerve combing” for trigeminal neuralgia without vascular compression: report of 10 cases. Clin J Pain. 2009;25(1):44–47.

24. Moher D, Liberati A, Tetzlaff J, Altman DG, Group P. Preferred reporting items for systematic reviews and meta-analyses: the PRISMA statement. J Clin Epidemiol. 2009;62(10):1006–1012.

25. Durnford AJ, Gaastra B, Akarca D, et al. Internal neurolysis: ‘nerve combing’ for trigeminal neuralgia without neurovascular conflict - early UK outcomes. Br J Neurosurg. 2020:1–4.

26. Hussain MA, Konteas A, Sunderland G, et al. Re-Exploration of Microvascular Decompression in Recurrent Trigeminal Neuralgia and Intraoperative Management Options. World Neurosurg. 2018;117:e67–e74.

27. Jie H, Xuanchen Z, Deheng L, et al. The long-term outcome of nerve combing for trigeminal neuralgia. Acta Neurochir (Wien). 2013;155(9):1703-1708; discussion 1707.

28. Liang X, Dong X, Zhao S, Ying X, Du Y, Yu W. A retrospective study of neurocombing for the treatment of trigeminal neuralgia without neurovascular compression. Ir J Med Sci. 2017;186(4):1033–1039.

29. Sabourin V, Mazza J, Garzon T, et al. Internal Neurolysis with and without Microvascular Decompression for Trigeminal Neuralgia: Case Series. World Neurosurg. 2020;143:e70–e77.

30. Wu M, Jiang X, Niu C, Fu X. Outcome of Internal Neurolysis for Trigeminal Neuralgia without Neurovascular Compression and Its Relationship with Intraoperative Trigeminocardiac Reflex. Stereotact Funct Neurosurg. 2018;96(5):305–310.

31. Wu M, Qiu J, Jiang X, et al. Diffusion tensor imaging reveals microstructural alteration of the trigeminal nerve root in classical trigeminal neuralgia without neurovascular compression and correlation with outcome after internal neurolysis. Magn Reson Imaging. 2020;71:37–44.

32. Zhang X, Xu L, Zhao H, et al. Long-Term Efficacy of Nerve Combing for Patients with Trigeminal Neuralgia and Failed Prior Microvascular Decompression. World Neurosurg. 2017;108:711–715.

33. Zhao H, Zhang X, Tang D, Li S. Nerve Combing for Trigeminal Neuralgia Without Vascular Compression. J Craniofac Surg. 2017;28(1):e15–e16.

34. Zhou X, Liu Y, Yue Z, Luan D, Zhang H, Han J. Comparison of nerve combing and percutaneous radiofrequency thermocoagulation in the treatment for idiopathic trigeminal neuralgia. Braz J Otorhinolaryngol. 2016;82(5):574–579.

35. Regis J, Tuleasca C, Resseguier N, et al. Long-term safety and efficacy of Gamma Knife surgery in classical trigeminal neuralgia: a 497-patient historical cohort study. Journal of neurosurgery. 2016;124(4):1079–1087.

36. Dandy WE. The Treatment of Trigeminal Neuralgia by the Cerebellar Route. Annals of surgery. 1932;96(4):787–795.

37. Zakrzewska JM, Lopez BC, Kim SE, Coakham HB. Patient reports of satisfaction after microvascular decompression and partial sensory rhizotomy for trigeminal neuralgia. Neurosurgery. 2005;56(6):1304–1311; discussion 1311-1302.

38. Piatt JH, Jr., Wilkins RH. Treatment of tic douloureux and hemifacial spasm by posterior fossa exploration: therapeutic implications of various neurovascular relationships. Neurosurgery. 1984;14(4):462–471.

39. Bederson JB, Wilson CB. Evaluation of microvascular decompression and partial sensory rhizotomy in 252 cases of trigeminal neuralgia. Journal of neurosurgery. 1989;71(3):359–367.

